# Sibling Control Analysis of Perinatal Health and Family Environment Factors Related to Childhood ADHD Symptoms

**DOI:** 10.1101/2025.06.16.25329516

**Authors:** Michael A. Mooney, Peter Ryabinin, Elizabeth Nousen, Jessica Tipsord, Nathan F. Dieckmann, Sarah L. Karalunas, Megan M. Herting, Molly Nikolas, Joel T. Nigg, Stephen V. Faraone

## Abstract

**Background:** Numerous studies have reported associations between environmental exposures and ADHD. However, whether environmental effects are causal or due to confounding with other familial factors, such as genetic risk, is still unclear. A more complete understanding of which environmental risk factors are causal remains crucial.

**Methods:** Using one population (ABCD cohort, N=11646) and one case-control cohort (Oregon ADHD-1000, N=744), we conducted both full-cohort and sibling-control analyses (770 and 152 families in ABCD and Oregon ADHD-1000, respectively) to assess the association of family environment and perinatal risk factors with ADHD symptoms. Within-family effects were compared to effects estimated in the full cohorts. We also assessed the impact of gene-environment correlation using child polygenic risk scores and measures of maternal mental health.

**Results:** For both cohorts, full-cohort analyses yielded significant associations between child ADHD symptoms and family conflict, perinatal health factors, and breastfeeding duration (p-values <0.001). These associations were non-significant after accounting for family-level confounds (e.g., genetic risk and shared family environment) in exposure-discordant sibling-control analyses. In the full cohorts, effect sizes were substantially reduced (an average 43.7% decrease in effect size across all exposures tested in the ABCD cohort; average 47.6% decrease in Oregon ADHD-1000) after adjusting for child ADHD polygenic risk and measures related to maternal mental health.

**Conclusions:** The full-cohort associations between child ADHD symptoms and environmental risks confirm associations from prior research, but current findings do not indicate direct causal effects. Instead, much or all of the observed risk appears due to confounding with family-level factors, likely including genetic factors. Our results underscore the importance of accounting for familial risk factors that may confound relationships between behavioral traits and environmental exposures by using multiple study designs.

**Key Points:**

- Numerous environmental risk factors are associated with ADHD, but whether these are important casual associations is insufficiently studied.
- Significant associations between child ADHD symptoms and features of the family environment and perinatal health were replicated in two independent cohorts, confirming prior work.
- Sibling-control analyses provided no consistent evidence of causal relationships for any of the risk factors studied. Furthermore, accounting for factors related to genetic risk and parental mental health substantially reduced the observed effect sizes in full-cohort analyses.
- Findings are consistent with previous studies on the effects of breastfeeding duration and some substance exposures (e.g., smoking during pregnancy), suggesting associations are largely due to confounding with unmeasured familial factors.
- Further study is warranted regarding potential sex differences in exposure associations with ADHD symptoms, as well as more complex causal pathways (e.g., gene-environment or environment-environment interaction).

## INTRODUCTION

Although twin studies (Faraone & Larsson, 2019) and large-scale genome-wide association studies (Demontis et al., 2023) have provided ample evidence that genetic liability plays an important role in the etiology of attention-deficit hyperactivity disorder (ADHD)—heritability estimates are 70-80% (Faraone & Larsson, 2019)—genetic factors alone do not explain the development and course of ADHD. Numerous environmental exposures have been associated with ADHD (Froehlich et al., 2011), yet studies often fail to consider genetic effects (i.e., gene-environment correlation) or other within-family confounders. A more precise understanding of the causality of environmental exposures is needed to fully understand ADHD etiology and promote effective prevention.

A recent systematic review of 63 meta-analyses examining 40 environmental exposures (Kim et al., 2020) found convincing or highly-suggestive evidence for associations between ADHD and several perinatal exposures: maternal acetaminophen use during pregnancy, maternal smoking during pregnancy, maternal hypertensive disorder during pregnancy, preeclampsia, and maternal pre-pregnancy obesity/overweight. Breastfeeding duration was the one protective factor significantly associated with ADHD, though the level of evidence was considered weak.

Still, breastfeeding duration has been a factor of considerable interest related to ADHD and other behavioral traits. A recent systematic review that focused on pregnancy-related factors and a meta-analysis found that ADHD was more common in the absence of any breastfeeding (Bitsko et al., 2024; Tseng et al., 2019). Associations with other mental health disorders, however, has been mixed (Bugaeva et al., 2023).

However, many of the studies examining risk or protective factors associated with mental health traits, such as ADHD, have been unable to assess causality. The recognition that many environmental exposures are correlated has prompted the use of study designs that control for potential confounders, including genetically-informed designs (Thapar & Rice, 2021). Sibling-control designs are able to control for factors shared within a family (e.g., genetic risk, parenting style, localized toxicant exposure) and therefore can distinguish between causal effects and effects due to confounding with unmeasured family-level factors (Begg & Parides, 2003; Frisell, 2021; Gustavson, Torvik, Davey Smith, Røysamb, & Eilertsen, 2024; Saunders, McGue, & Malone, 2019; Sjölander, Frisell, & Öberg, 2022).

For example, as stated above, an association between ADHD and maternal smoking during pregnancy is well replicated (Dong et al., 2018; Huang et al., 2018). Yet studies that controlled for possible confounders, reported that this association is likely due to confounding with genetic risk or other family-level factors (D’Onofrio et al., 2008; Gustavson et al., 2017; Thapar & Rice, 2021). Likewise, apparent associations between ADHD and prenatal acetaminophen exposure (Gustavson et al., 2021) as well as breastfeeding duration (Colen & Ramey, 2014; Der, Batty, & Deary, 2006) have been shown to be due to confounding with other, unmeasured family-level factors. Indeed, Nigg *et al*. reported a reverse causality in which child ADHD led to reduced breastfeeding duration (presumably due to precursive developmental difficulties, though genetic risk was not examined in that study) (Nigg, Stadler, von Eye, & Wiedermann, 2020).

The influence of the family environment on ADHD symptoms has also been an area of significant interest, and studies have consistently found associations between ADHD symptoms and parenting factors (Claussen et al., 2024), as well as family conflict (Pressman et al., 2006; Sharp, Mangalmurti, Hall, Choudhury, & Shaw, 2021). Improved parenting skills have been shown to be important in successful ADHD treatment in randomized trials (Doffer et al., 2023).

However, like other exposures, whether effects of the family environment on ADHD are causal is not well established. For instance, recent studies have suggested that the effects of household chaos are at least partly due to gene-environment correlation (Agnew-Blais et al., 2022), and that changes in the family environment, such as increased conflict, follow (rather than cause) increases in ADHD symptoms (Brinksma et al., 2020; Saulsbury et al., 2025; Selah et al., 2024). There is evidence, however, that family processes may be important mediators of changes in ADHD symptoms (e.g., parenting processes influencing treatment response) (Hinshaw, Arnold, & For the MTA Cooperative Group, 2015).

Investigating to what extent environmental risk factors are causal is crucial for understanding etiology and informing risk assessment and interventions. For these reasons, prior studies point to the importance of utilizing multiple study designs to explore the complex relationships between environmental factors and mental health (D’Onofrio et al., 2016; Frisell, 2021; Thapar & Rice, 2021).

The first goal of the current study was to use a sibling-control study design, in two independent cohorts, to investigate the effects of selected environmental exposures on childhood ADHD symptoms and to assess the role of unmeasured familial confounds. Using data from the Adolescent Brain Cognitive Development (ABCD) Study (Jernigan, Brown, & Dowling, 2018; Volkow et al., 2018) and the Oregon ADHD-1000 cohort (Nigg et al., 2023), we studied two measures of the family environment (stressful life events and family conflict), four factors related to mother and child perinatal health (prenatal exposures, maternal health during pregnancy, birth characteristics, and infant health), and breastfeeding duration. The second goal was to investigate the impact of measured family-level factors (genetic risk, parental mental health status, and neighborhood disadvantage), which are likely to be confounded with the other environmental exposures of interest.

## MATERIALS AND METHODS

### Participants

Primary analyses were conducted using data from the Adolescent Brain Cognitive Development (ABCD) Study cohort (release 5.1; http://dx.doi.org/10.15154/z563-zd24). The ABCD cohort is a large, 21-site, diverse sample of children aged 9-10 years at baseline (Jernigan et al., 2018; Volkow et al., 2018). Analyses were conducted both in the full ABCD cohort, as well as a matched ADHD case-control subsample. ADHD cases (N=411) were identified based on the Tier 4 criteria previously reported by Cordova *et al*. (Cordova et al., 2022). Non-ADHD controls were selected at random, but were matched on sex assigned at birth, age, self-reported race/ethnicity, and study site (N=820 controls) (Mooney et al., 2023). Replication analyses were conducted in the Oregon ADHD-1000 cohort, a community recruited case-control cohort of youth age 7-11 years at baseline that is enriched with ADHD cases (Nigg et al., 2023). All analyses were limited to subjects with genotype data. Thus, for this report the full cohort sample size was 11646 for the ABCD cohort and 744 for the Oregon ADHD-1000.

For the sibling-control analyses, siblings in both cohorts were confirmed genetically, using the inference criteria for full siblings given in Equation 1 (Manichaikul et al., 2010), and by self-report, resulting in 770 families in the ABCD cohort and 152 families in the Oregon ADHD-1000 cohort. A small fraction of families included in the sibling analyses had more than two siblings (5% in ABCD; 2% in Oregon ADHD-1000). Due to missing data and the requirement for siblings to have discordant exposures, the sample sizes varied across analyses and are reported in results tables.

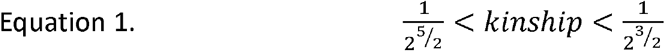

### ADHD Symptom Measures

Measures of ADHD symptoms were the outcomes for all analyses. In the ABCD cohort, T-scores from the CBCL ADHD DSM5-oriented scale were used (Achenbach & Rescorla, 2001). In the Oregon ADHD-1000 cohort, inattention T-scores from the Conners (3^rd^ edition) were used (Conners, 2008). Sensitivity analyses were done to confirm effects on additional measures of ADHD symptoms in the Oregon ADHD-1000 (total symptoms T-score from the ADHD Rating Scale and inattention raw scores from the Conners; see Supplemental Materials). We chose not to examine ADHD diagnosis given the very small number of ADHD cases among siblings in the ABCD cohort.

### Environmental Exposures

Environmental exposures were selected such that a minimum of 100 subjects (50 sibling pairs) were available for the discordant sibling analyses. In both cohorts, family conflict was measured using the conflict subscale from the family environment scale (FES) (Moos & Moos, 2002). Stressful life events was measured in the ABCD cohort by a sum score from the parent-reported life events (LE) scale at the 1-year follow-up (Barch et al., 2018; Hoffman et al., 2019), and in the Oregon ADHD-1000 by a sum score of the parental stress index (PSI) stressful life events scale (Ríos, Zekri, Alonso-Esteban, & Navarro-Pardo, 2022). Breastfeeding duration was measured as number of months after birth in the ABCD cohort, but was reported ordinally in the Oregon cohort using the following scale: 0=0 months, 1=1-3 months,2= 4-6 months, 3=7-12 months, 4=13-18 months, and 5=19 months or more.

Factor analyses were conducted to generate four reliable indices of perinatal exposures: prenatal substance exposures, maternal health during pregnancy, birth complications, and infant perinatal health. In each case, a higher score indicates a greater adverse exposure. The confirmatory factor analyses included 39 indicators from the ABCD cohort and 35 indicators from the Oregon cohort, and a four-factor model fit well in both cohorts (TLI/CFI >0.92 and RSMEA = 0.02 in ABCD; TLI/CFI >0.86 and RSMEA = 0.03 in Oregon ADHD-1000). The code for generating these factor scores in ABCD is publicly available (https://osf.io/76tqa) (Nikolas, Rounds, Shearer, & Mooney, 2024).

Thus, in total, 7 exposure variables were studied: the 4 perinatal factor scores, family conflict, stressful events, and breastfeeding duration.

### Statistical Analyses – Full Cohorts

Linear mixed-effect models were used to examine the association between each exposure and ADHD symptoms in both the full cohorts. Models were implemented using the lmerTest R package (Kuznetsova, Brockhoff, & Christensen, 2017). Models included covariates for age, sex assigned at birth, and the first three genetic principal components (to control for potential effects of ancestry), as well as a random effect for family to account for nesting. In line with recommendations, a random effect for study site was also included for models run in the ABCD cohort. All exposures and ADHD measures were scaled to have a mean of 0 and standard deviation of 1 prior to analysis.

Because seven exposure variables were studied, a conservative corrected p-value threshold of 0.05/7=0.007 was used to determine statistical significance.

### Statistical Analyses: Exposure-discordant Siblings

Sibling-control analyses were done only among siblings discordant for the exposure of interest, and included a random effect for family and covariates for age and sex. The sibling-control analyses estimated both within- and between-family effects by also including a term for the mean family-level exposure, as described in (Begg & Parides, 2003). These models partition the exposure effect into individual-level (within-family) and group-level (between-family) effects, with the individual-level effect representing a causal effect after adjusting for family-level confounders. Hence, a lack of significant within-family effect suggests the presence of confounding.

### Sensitivity Analyses: Full Cohorts

In the full cohorts, sensitivity analyses were performed to control for measured family-level factors, which may partially explain the observed effects of exposures due to confounding. These adjusted analyses included the following covariates: child ADHD polygenic risk score (details in Supplemental Materials), maternal age at birth, highest parental education, household income, and measures of parental mental health symptoms. In the ABCD cohort, parental mental health was measured by the Adult Self Report (ASR) ADHD DSM-oriented T-score (Achenbach & Rescorla, 2003), a binary indicator of maternal lifetime depression problems (family history module), and the Personal Strengths ASR adaptive functioning scale T-score (Achenbach & Rescorla, 2003). In the Oregon ADHD-1000, the following measures were used: maternal diagnoses for ADHD and major depressive disorder from the Kiddie Schedule of Affective Disorders and Schizophrenia (KSADS), and maternal clinician-administered Structured Clinical Interview for DSM-IV global assessment of functioning (GAF) score (Pedersen, Urnes, Hummelen, Wilberg, & Kvarstein, 2018). The following covariates were included to control for other environmental exposures: child PTSD diagnosis (as a proxy for trauma) from the KSADS, as well as two residentially derived census estimates for neighborhood disadvantage: the child opportunity index (COI) (https://www.diversitydatakids.org/child-opportunity-index) and the area deprivation index (ADI) (Singh, 2003). For the COI, the three subscales (educational, health and environment, social-economic) and the total score were used (Ferrara et al., 2024). In the Oregon ADHD-1000 cohort, participants with visits prior to 2012 were assigned COI values based on COI 2.0, otherwise COI values were based on COI 3.0.

### Sensitivity Analyses: Exposure-discordant Siblings

To examine the potential for different environmental effects between sexes, sibling analyses (using the same methods as the primary sibling analyses described above) were conducted separately among families with male-male sibling pairs and those with female-female sibling pairs.

In addition, several sensitivity analyses were conducted to examine the impact of known sources of bias in sibling-control analyses (Gustavson et al., 2021). First, the impact of measurement error was estimated using simulations implemented in the SibSim tool (https://kristin-gustavson.shinyapps.io/sibsimextended/; https://osf.io/ryszk/) described in (Gustavson et al., 2024). Second, the impact of non-shared confounders was assessed by calculating between-sibling correlation of measured confounders and comparing those to the between-sibling correlation of exposures of interest. Finally, the impact of carry-over effects was assessed in two ways: (1) by testing whether the younger sibling’s exposure level was associated with the older sibling’s ADHD symptoms, while controlling for the older sibling’s exposure, and (2) by stratifying families in the sibling subsample depending on whether the older or younger sibling has the higher exposure and then comparing effect sizes between those two groups of families (Sjölander, Frisell, Kuja-Halkola, Öberg, & Zetterqvist, 2016). The analyses assessing carry-over effects were limited to families with two siblings only.

## RESULTS

### Description of cohorts

Overviews of each cohort and the discordant-sibling subsamples are provided in Tables 1 and 2. Due to different study designs, there was a difference in the distribution of ADHD symptoms between the ABCD and Oregon full cohorts (p <0.001). The average CBCL ADHD DSM-oriented T-score was 53.2 in the ABCD cohort reflecting community averages, while the average Conners 3 inattention T-score was 64.6 in the Oregon ADHD-1000 cohort, reflecting the over-sampling of ADHD cases. Though the mean age was similar between the two cohorts, due to the design differences, the cohorts differed in age range (9 to 11 years in ABCD; 7 to 13 years in Oregon ADHD-1000), as well as sibling age difference (mean difference of 1.27 and 1.97 years in ABCD and Oregon ADHD-1000, respectively; p <0.001).

**Table 1.** Overview of cohorts. For continuous measures mean and standard deviation (in parentheses) are reported. *In the ABCD cohort, “other” was a distinct race/ethnicity category. In the Oregon ADHD-1000 cohort, “other” encompassed Native American/Alaska Native/Eskimo, Native Hawaiian/Pacific Islander, or self-reported “other”. **ADHD symptoms were measured by the CBCL ADHD DSM-oriented T-score in the ABCD cohort and the Conners 3 inattention T-score in the Oregon ADHD-1000.

|  | ABCD | Oregon ADHD-1000 |
| --- | --- | --- |
| Participants | 11646 | 744 |
| Age in years | 9.48 (0.0508) | 9.44 (1.54) |
| % Female | 47.9% | 38.4% |
| Race / Ethnicity |  |  |
| White, non-Hispanic | 6072 | 606 |
| Hispanic / Latino/a | 2353 | 34 |
| Black / African-American | 1747 | 49 |
| Asian | 249 | 35 |
| Other / Unknown* | 1225 | 19 |
| ADHD T-score** | 53.2 (5.63) | 64.6 (16.8) |
| Family Income | Median = \$75,000 - \$99,999 | Median = \$75,000 - \$99,999 |
| Family Conflict | 2.54 (1.96) | 2.84 (2.10) |
| Parental Stress | 3.17 (2.62) PLE scale | 1.23 (1.53) PSI scale |
| Maternal Health Score | 0.106 (0.674) | 0.112 (0.634) |
| Prenatal Exposure Score | 0.113 (0.669) | 0.133 (0.629) |
| Infant Health Score | 0.0945 (0.657) | 0.0836 (0.589) |
| Birth Characteristics Score | 0.0642 (0.808) | 0.0126 (0.776) |
| Breastfeeding duration | 7.77 (8.75) months | Median = 7-12 months |

**Table 2.** Discordant-sibling subsamples. *Indicates a significant difference compared to the corresponding full cohort (p<0.05).

|  | ABCD | Oregon ADHD-1000 |
| --- | --- | --- |
| Family size: No. families | 2: 730, 3: 38, 4: 1, 5: 1 | 2: 149, 3: 3 |
| Age in years | 9.44 (0.0507)* | 9.54 (1.69) |
| % Female | 48.8% | 43.0% |
| Race/Ethnicity |  |  |
| White, non-Hispanic | 935 | 254 |
| Hispanic / Latino/a | 286 | 19 |
| Black / African-American | 162 | 16 |
| Asian | 30 | 10 |
| Other / Unknown | 172 | 8 |
| ADHD T-score | 52.6 (5.00)* | 58.6 (16.0)* |
| Family Income | Median = \$75,000 - \$99,999 | Median = \$75,000 - \$99,999 |
| Family Conflict | 2.42 (2.05) | 3.11 (2.23) |
| Parental Stress | 3.07 (2.69)* PLE scale | 1.38 (1.51) PSI scale |
| Maternal Health Score | -0.0722 (0.595)* | 0.0453 (0.593) |
| Prenatal Exposure Score | 0.00145 (0.601)* | 0.0206 (0.489) |
| Infant Health Score | -0.0498 (0.585)* | 0.0417 (0.551) |
| Birth Characteristics Score | -0.132 (0.651)* | 0.00 (0.755) |
| Breastfeeding duration | 8.29 (8.06) months* | Median = 7-12 months |

In the ABCD cohort, the discordant-sibling subsample had significantly lower ADHD symptoms and lower adverse exposure levels on average (Table 2). In the Oregon ADHD-1000 cohort, ADHD symptoms were significantly lower in the sibling subsample compared to the full cohort, but there were no significant differences in exposure levels.

A description of the ABCD matched ADHD case-control subsample is reported in Supplemental Table S1.

### Exposure associations in the full cohorts

In the ABCD cohort, family conflict, stressful life events score, duration of breastfeeding and all four perinatal health factors were significantly associated with childhood ADHD symptoms. All significant associations, except for the birth complication factor, replicated in the Oregon ADHD-1000. However, the effect of stressful events did not survive multiple testing correction in the replication cohort (Table 3).

**Table 3.** Associations with ADHD symptoms in the full cohorts. N=total sample size analyzed. Exposures in bold were statistically significant in both cohorts. Regression models included covariates for age, sex, and the first three genomic principal components. Random effects were included for family and study site (for ABCD only).

|  | ABCD |  | Oregon ADHD-1000 |  |
| --- | --- | --- | --- | --- |
| | $\beta$ (SE) | P-value (N) | $\beta$ (SE) | P-value (N) |
| **Family Conflict** | **0.199 (0.009)** | **4.67e-97 (11646)** | **0.138 (0.037)** | **2.25e-04 (720)** |
| Stress | 0.162 (0.010) | 2.24e-61 (11028) | 0.085 (0.040) | 0.036 (646) |
| **Maternal Health Prob.** | **0.148 (0.010)** | **3.75e-51 (11600)** | **0.183 (0.036)** | **6.08e-07 (739)** |
| **Prenatal Exposures** | **0.175 (0.010)** | **2.71e-74 (11600)** | **0.177 (0.036)** | **1.41e-06 (735)** |
| **Infant Health Prob.** | **0.084 (0.010)** | **6.65e-18 (11600)** | **0.144 (0.036)** | **8.99e-05 (735)** |
| Birth Characteristics | 0.052 (0.010) | 4.46e-07 (11600) | 0.041 (0.037) | 0.267 (735) |
| **Breastfeeding Dur.** | **-0.051 (0.010)** | **2.82e-07 (11222)** | **-0.186 (0.037)** | **4.95e-07 (735)** |

Effect estimates in the ABCD matched ADHD case-control subsample were stronger than in the full ABCD cohort but were otherwise consistent (Supplemental Table S2).

Effect estimates when using alternate measures of ADHD symptoms available in the Oregon ADHD-1000 are provided in Supplemental Table S3 and are consistent with effects observed in the primary analysis.

### Sensitivity analyses: Adjusting for measured confounders in the full cohorts

In the full ABCD cohort, accounting for child ADHD genetic risk and factors related to parental mental health substantially reduced the effects of environmental exposures on child ADHD symptoms (Table 4). Family conflict, stressful life events, maternal health during pregnancy, prenatal substance exposures, infant health, and breastfeeding duration all remained significantly associated with child ADHD symptoms in the adjusted models, but the reduction in effect sizes ranged from 27.2% for breastfeeding duration to 51.9% for stressful life events. The effect of birth complications, which did not replicate in the unadjusted analyses, dropped 49.1% and was only nominally significant after adjustment.

**Table 4.** Full-cohort analyses adjusted for child polygenic risk for ADHD and potential confounders related to parent mental health. Exposures in bold were statistically significant in both cohorts. Regression models included covariates for age, sex, child polygenic risk for ADHD, maternal age at birth, highest parental education, household income, parent ADHD symptoms, parent depression problems, and parent GAF score.

|  | ABCD |  | Oregon ADHD-1000 |  |
| --- | --- | --- | --- | --- |
| | $\beta$ (SE) | P-value (N) | $\beta$ (SE) | P-value (N) |
| **Family Conflict** | **0.119 (0.010)** | **1.87e-32 (9937)** | **0.143 (0.042)** | **7.0e-4 (542)** |
| Stress | 0.078 (0.010) | 8.00e-14 (9463) | 0.023 (0.047) | 0.628 (490) |
| Maternal Health Prob. | 0.081 (0.010) | 1.19e-15 (9937) | 0.106 (0.043) | 0.014 (561) |
| Prenatal Exposures | 0.093 (0.012) | 4.30e-20 (9937) | 0.115 (0.043) | 0.009 (561) |
| Infant Health Prob. | 0.045 (0.012) | 5.80e-06 (9937) | 0.066 (0.042) | 0.114 (561) |
| Birth Complications | 0.026 (0.012) | 0.011 (9937) | 0.001 (0.040) | 0.984 (561) |
| **Breastfeeding Dur.** | **-0.037 (0.012)** | **2.62e-04 (9734)** | **-0.121 (0.043)** | **0.005 (559)** |

Likewise, in the ABCD matched case-control subsample, exposure effect estimates in adjusted models were reduced by 44.7% on average (Table S2).

In the full Oregon ADHD-1000 cohort, a similar pattern of results was seen in adjusted models (Table 5), except for family conflict, where the effect size increased slightly (3.6% increase). In addition to family conflict, breastfeeding duration remained statistically significant after adjustment for covariates, but the effect size dropped 34.9%. The effects of maternal health and prenatal exposures were reduced by 42.1% and 35.0%, respectively, and were only nominally significant after adjustment. Stressful life events, infant health, and birth complications were not significant in the covariate-adjusted models.

**Table 5.**
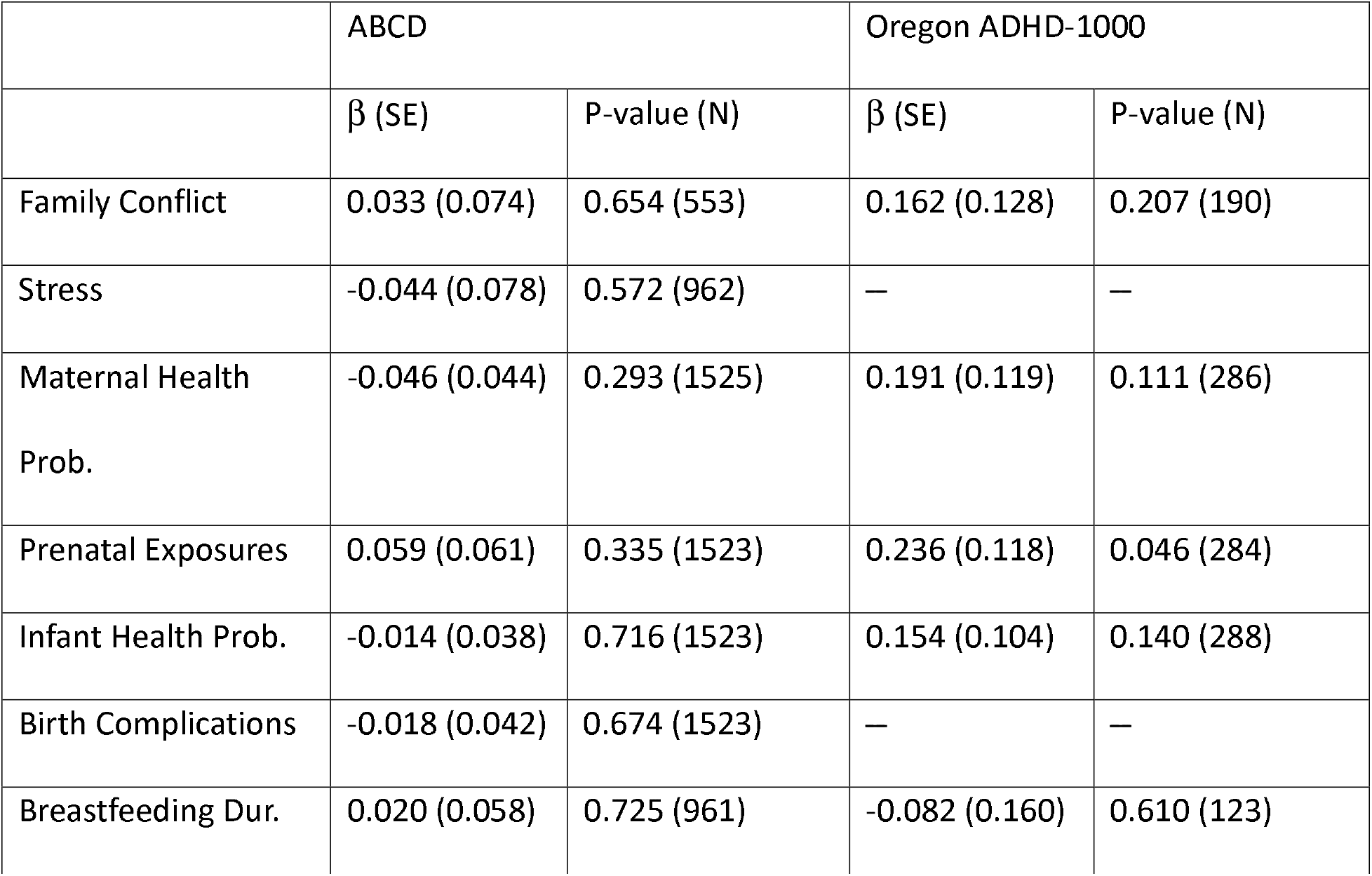
Associations with ADHD symptoms in the discordant-sibling subsamples. Within-family effects are shown here. Results are shown for only those exposures with significant effects in the full cohorts. Regression models included covariates for age, sex, and the mean family-level exposure, as well as random effects for family and study site (ABCD only).

|  | ABCD |  | Oregon ADHD-1000 |  |
| --- | --- | --- | --- | --- |
| | $\beta$ (SE) | P-value (N) | $\beta$ (SE) | P-value (N) |
| Family Conflict | 0.033 (0.074) | 0.654 (553) | 0.162 (0.128) | 0.207 (190) |
| Stress | -0.044 (0.078) | 0.572 (962) | — | — |
| Maternal Health Prob. | -0.046 (0.044) | 0.293 (1525) | 0.191 (0.119) | 0.111 (286) |
| Prenatal Exposures | 0.059 (0.061) | 0.335 (1523) | 0.236 (0.118) | 0.046 (284) |
| Infant Health Prob. | -0.014 (0.038) | 0.716 (1523) | 0.154 (0.104) | 0.140 (288) |
| Birth Complications | -0.018 (0.042) | 0.674 (1523) | — | — |
| Breastfeeding Dur. | 0.020 (0.058) | 0.725 (961) | -0.082 (0.160) | 0.610 (123) |

Adjusting for measures of neighborhood disadvantage also reduced effect sizes, though to a lesser extent than seen for genetic risk and parental mental health (Supplemental Table S6).

### Between- and within-family effects

To assess the impact of genetic and familial confounding on exposure associations, those showing significant associations in the full cohorts were tested in the discordant-sibling subsamples. In the ABCD cohort, for all exposures, between-family effects in the discordant-sibling subsample were consistent with effects seen in the full cohort, though only effects for the maternal and infant health perinatal factors were significant after multiple-testing correction (Supplemental Table S4). No within-family effects were statistically significant (Table 5), suggesting associations in the full cohort are likely due to confounding with family-level factors.

In the Oregon ADHD-1000 discordant-sibling subsample, none of the between-family nor within-family effects were significant after multiple testing correction (Table 5 and Supplemental Table S5). The within-family effect of prenatal substance exposures, however, was nominally significant.

### Sensitivity analyses: Sex differences and sources of bias in sibling-control studies

In the ABCD cohort, sibling analyses stratified by sex identified a statistically significant within-family effect for prenatal substance exposures among families with female-only sibling pairs. As in the primary analyses, no significant within-family effects were observed among families with male-only sibling pairs (Supplemental Table S8).

Additional sensitivity analyses examining various potential sources of bias in sibling studies showed: (1) Only a small potential impact of measurement error on the findings reported here. Specifically, moderate measurement error may explain the reduction in effect size for prenatal substance exposure between the full-cohort model and the sibling-control model (Supplemental Table S9). (2) In the ABCD cohort, there was some evidence of carry-over effects of the older sibling’s ADHD symptoms on the younger sibling’s maternal health and birth characteristics factors, though it is unlikely that the magnitude of carry-over effects observed would explain the findings reported (Supplemental Table S11). (3) The risk of bias due to non-shared confounders appears low in both cohorts (Supplemental Table S14 and S15).

Further details on all sensitivity analyses are presented in the Supplemental Materials.

## DISCUSSION

Once risk factors are identified for ADHD (and other conditions), determining their causal importance is the next step. Sibling-control analyses are one way to estimate such effects and were employed here. As expected, we identified replicable associations between ADHD symptoms and family conflict, prenatal substance exposures, maternal health during pregnancy, infant health, and breastfeeding duration. However, in sibling-control analyses there was no consistent evidence of within-family effects for any of the exposures examined, suggesting that (1) any causal associations were too small to detect here and/or (2) these patterns of associations are largely due to confounding with family-level factors (including genetic risk). This interpretation was further supported by the observation that adjusting for child polygenic risk of ADHD and potential family-level confounders, such as factors related to parental mental health, substantially reduced the strength and significance of the observed associations in both full cohorts.

It is important to note that this analysis does not rule out all possible mechanistic connections between environmental risk factors and ADHD. For example, genotype-by-environment and environment-by-environment interaction effects were not examined here. Nor do our findings rule out the potential importance of family environment as a mediator of other causal factors (Johnston & Mash, 2001). Furthermore, while we used child polygenic scores and measures of maternal mental health to assess the impact of gene-environment correlation, directly measuring parent genotypes would better enable disentangling direct genetic effects from indirect genetic effects (genetic nurture) (Pingault et al., 2023).

Nonetheless, these findings are important to help narrow the scope of targets for early intervention to improve outcomes for very young children at risk for ADHD. Results suggest that breastfeeding duration is not causally related to ADHD symptoms in children. Indeed, Nigg et al. suspected a reverse causality (that ADHD or its precursive developmental factors contribute to reduced breastfeeding duration) (Nigg et al., 2020). Findings here are consistent with prior studies that examined the relationship between breastfeeding and other behavioral traits (Colen & Ramey, 2014; Der et al., 2006; Jallow, Hurtig, Kerkelä, Miettunen, & Halt, 2024) suggesting limited or no causal effects. Whether extended breastfeeding could still protect at-risk infants from other unmeasured exposures, which in turn could contribute to exacerbated ADHD symptoms, would be a valuable next investigation.

Likewise, our failure to detect causal effects between ADHD symptoms and perinatal factors or family conflict measures are consistent with prior studies using both family-based designs and other methods for adjusting for possible confounders (Agnew-Blais et al., 2022; Carlsson, Molander, Taylor, Jonsson, & Bölte, 2021; Curran et al., 2015; D’Onofrio et al., 2008; Gustavson et al., 2017, 2021; Lemelin, Sheehy, Zhao, & Bérard, 2021; Saulsbury et al., 2025).

Our findings suggest potential sex differences for both between- and within-family exposure effects, though we were unable to test generalizability of these differences. Sex differences among environmental influences on behavioral traits should be a focus of further study (Momany et al., 2017; Momany, Jasper, Markon, Nikolas, & Ryckman, 2023).

In contrast to our findings, a recent analysis of cumulative adverse exposures during the perinatal period, conducted in the ABCD cohort, found that the difference in cumulative exposure between siblings was significantly associated with difference in CBCL total problems between siblings (Roffman et al., 2021). It should be noted, however, that this within-family effect was substantially smaller (*β*=0.67, p=0.017) than the covariate-adjusted effect observed in a non-sibling sample (*β*=1.94, p<0.001).

Several potential sources of bias in sibling-control analyses were examined in the current study. Both measurement error and carry-over effects were found to have a potential impact on findings reported here. Measurement error is certainly a concern, as multiple factors examined here relied on parent recall years after the child’s birth. Studies suggest very good maternal recall of breastfeeding duration, even up to 20 years after the child’s birth (Li, Ingol, Smith, Oza-Frank, & Keim, 2020; Natland, Andersen, Nilsen, Forsmo, & Jacobsen, 2012), while estimates of recall reliability for other pregnancy-related events has been mixed (Chin et al., 2017; Tomeo et al., 1999; Yawn, Suman, & Jacobsen, 1998).

Carry-over effects related to maternal perinatal health and birth characteristics may have biased the effects reported, though it is unlikely the amount of bias introduced would change the overall conclusions drawn here (Sjölander et al., 2016). Non-shared confounders were unlikely to have biased the observed effects.

There are several limitations that call for additional research. First, the ABCD cohort, being a community sample, has only a small proportion of participants who met our criteria for ADHD. The restricted range of symptoms across the full cohort likely attenuated effect estimates and may have prevented detection of effects that are seen only among individuals with elevated symptoms. Further, the limited age range of ABCD participants (ages 9-10 at baseline) resulted in limited within-family variability among siblings, particularly for exposures related to the family environment, which in turn substantially limited the power to detect within-family effects for those exposures. Finally, neither cohort was recruited with regard to perinatal or other environmental exposures.

While the different ascertainment strategy (i.e., the enrichment of ADHD cases) and wider age range of the Oregon ADHD-1000 cohort provided a good test of generalizability, the small sample size for the sibling analysis in the Oregon ADHD-1000 was a notable limitation in detecting smaller but potentially important effects. These differences may explain the different pattern of effects observed (larger within-family effect sizes) in the Oregon ADHD-1000. However, given the small sample size, and only nominal significance, these results should be interpreted cautiously.

In addition, we were unable to perform a sibling analysis within the matched ADHD case-control subsample of ABCD, due to the small number of siblings among cases. Further study in samples enriched for ADHD cases will be important to determine whether environmental effects may be different in individuals at higher risk of ADHD or with more severe exposures (Kennedy et al., 2016).

Finally, while not necessarily a limitation, it is important to recognize the difference in exposure timing between the perinatal and family environment exposures studied here, with measures of family environment collected at the same time as ADHD symptom assessment. Longitudinal studies will be important for determining transient versus long-lasting effects (Livingstone et al., 2016), as well as how correlations between environmental exposures and other risk factors vary over time.

Overall, our findings are consistent with prior genetically informed studies on the effects of certain environmental risk factors on child ADHD symptoms and support the importance of accounting for genetic risk and other family-level factors when examining environmental risks for ADHD. Our work and others’ suggest that cohort studies of environmental exposures, without adequate controls for potential confounding, should be interpreted with caution with regard to causality (Faraone, 2024). It would seem premature for any such studies to be interpreted as evidence for the necessity of preventive measures. At the same time, further study of more complex chains of effect, such as interaction effects, remain important to fully understand the role of environmental risk factors in ADHD.

## Data Availability

This study involves publicly available data, which can be obtained from the NIMH Data Archive (NDA Study 2313 and NDA Collection 2857).

http://dx.doi.org/10.15154/z563-zd24

https://nda.nih.gov/edit_collection.html?id=2857

## ACKNOWLEDGEMENTS

Data used in the preparation of this article were obtained from the Adolescent Brain Cognitive Development^SM^ (ABCD) Study (https://abcdstudy.org), held in the NIMH Data Archive (NDA). This is a multisite, longitudinal study designed to recruit more than 10,000 children age 9-10 and follow them over 10 years into early adulthood. The ABCD Study^®^ is supported by the National Institutes of Health and additional federal partners under award numbers U01DA041048, U01DA050989, U01DA051016, U01DA041022, U01DA051018, U01DA051037, U01DA050987, U01DA041174, U01DA041106, U01DA041117, U01DA041028, U01DA041134, U01DA050988, U01DA051039, U01DA041156, U01DA041025, U01DA041120, U01DA051038, U01DA041148, U01DA041093, U01DA041089, U24DA041123, U24DA041147. A full list of supporters is available at https://abcdstudy.org/federal-partners.html. A listing of participating sites and a complete listing of the study investigators can be found at https://abcdstudy.org/consortium_members/. ABCD consortium investigators designed and implemented the study and/or provided data but did not necessarily participate in the analysis or writing of this report. This manuscript reflects the views of the authors and may not reflect the opinions or views of the NIH or ABCD consortium investigators. The ABCD data repository grows and changes over time. The ABCD data used in this report came from the ABCD 5.1 data release (http://dx.doi.org/10.15154/z563-zd24). Additional support for this work was made possible from NIEHS R01-ES032295 and R01-ES031074.

This work was supported by the National Institute of Mental Health of the National Institutes of Health under award numbers R01MH131685 and R37MH59105. The project was also supported by the National Center for Advancing Translational Sciences (NCATS), National Institutes of Health, through grant award number UL1TR002369. The content is solely the responsibility of the authors and does not necessarily represent the official views of the National Institutes of Health.

## FINANCIAL DISCLOSURES

In the past year, Dr. Faraone received income, potential income, travel expenses, continuing education support, and/or research support from Aardvark, Aardwolf, Tris, Otsuka, Ironshore, Johnson & Johnson/Kenvue, Corium, Supernus, Sky Therapeutics, Mentavi, and Genomind. With his institution, he has US patent US20130217707 A1 for the use of sodium-hydrogen exchange inhibitors in the treatment of ADHD. He also receives royalties from books. His educational websites (www.ADHDinAdults.com; www.ADHDevidence.org) are supported by The Upstate Foundation, Corium, Tris, Otsuka, and Supernus.

All other authors have declared no competing interests.

## Notes

### Funding Statement

Data used in the preparation of this article were obtained from the Adolescent Brain Cognitive Development^SM^ (ABCD) Study (https://abcdstudy.org), held in the NIMH Data Archive (NDA). This is a multisite, longitudinal study designed to recruit more than 10,000 children age 9-10 and follow them over 10 years into early adulthood. The ABCD Study® is supported by the National Institutes of Health and additional federal partners under award numbers U01DA041048, U01DA050989, U01DA051016, U01DA041022, U01DA051018, U01DA051037, U01DA050987, U01DA041174, U01DA041106, U01DA041117, U01DA041028, U01DA041134, U01DA050988, U01DA051039, U01DA041156, U01DA041025, U01DA041120, U01DA051038, U01DA041148, U01DA041093, U01DA041089, U24DA041123, U24DA041147. A full list of supporters is available at https://abcdstudy.org/federal-partners.html. A listing of participating sites and a complete listing of the study investigators can be found at https://abcdstudy.org/consortium_members/. ABCD consortium investigators designed and implemented the study and/or provided data but did not necessarily participate in the analysis or writing of this report. This manuscript reflects the views of the authors and may not reflect the opinions or views of the NIH or ABCD consortium investigators. The ABCD data repository grows and changes over time. The ABCD data used in this report came from the ABCD 5.1 data release (http://dx.doi.org/10.15154/z563-zd24). Additional support for this work was made possible from NIEHS R01-ES032295 and R01-ES031074.
This work was supported by the National Institute of Mental Health of the National Institutes of Health under award numbers R01MH131685 and R37MH59105. The project was also supported by the National Center for Advancing Translational Sciences (NCATS), National Institutes of Health, through grant award number UL1TR002369. The content is solely the responsibility of the authors and does not necessarily represent the official views of the National Institutes of Health.

### Author Declarations

IRB of Oregon Health & Science University gave ethical approval for this work.

