## Supplemental Materials for "Sibling Control Analysis of Perinatal Health and Family Environment Factors Related to Childhood ADHD Symptoms"

### METHODS:

#### *Polygenic risk scores:*

ADHD polygenic risk scores, in both cohorts, were calculated with the PRS-CSx tool (Ge et al., 2019; Ruan et al., 2022) using the ADHD-2022 PGC-iPSYCH genome-wide association study (Demontis et al., 2023) as the discovery data set and linkage disequilibrium reference panels from the 1000 Genomes phase 3. For the ABCD cohort, imputed genotypes provided in the ABCD 5.1 data release (http://dx.doi.org/10.15154/z563-zd24) were used. In the Oregon ADHD-1000, genotypes were imputed using the 1000 Genomes phase 3 reference panel as described previously (Nigg et al., 2018). PLINK (Chang et al., 2015) was subsequently used to calculate individual polygenic risk scores as the dot product of the SNP weights (output from PRS-CSx) and genotypes.

*Matched ADHD case-control sub-sample of the ABCD cohort:*

Uncontrolled (full cohort) association analyses were also run in a matched ADHD case-control sub-sample of ABCD, which consisted of N=411 putative ADHD cases and N=820 matched controls (Table S1). Results from these analyses are presented in Table S2.

*ADHD Symptom Measures:*

Additional ADHD symptom measures were tested in the Oregon ADHD-1000 full cohort analyses, namely parent-reported ADHD Rating Scale total symptoms T-score and parent-reported Conners 3 hyperactivity T-score. Associations were consistent across all symptom measures examined (Table S3).

*Assessing the impact of measurement error:*

The SibSim tool (https://kristin-gustavson.shinyapps.io/sibsimextended/) was used to simulate the impact of different levels of exposure measurement reliability, assuming that a true causal effect exists. Causal effect sizes for each exposure were assumed to be roughly equivalent to the effect size estimated in the ABCD full cohort analyses (an effect size of 0.1 was used for all exposures with estimated effects < 0.1). The number of families was set to N=700 and the percentage of families with more than two siblings was set to 1% for all simulations. Measurement reliability was varied between 0.7 and 0.9 for all exposures (for those with high observed correlations [>0.8], a reliability of 0.95 was also used). Results from these simulations are presented in Tables S9 and S10.

*Assessing the impact of carry-over effects:*

Carry-over effects occur when one sibling's exposure/outcome influences the other sibling's exposure/outcome. These effects have the potential to bias effect estimates in a sibling control analysis. Two types of potential carry-over effects were examined in the present study: (1) the effect of the older sibling's ADHD symptoms on the younger sibling's exposure level, and (2) the effect of the older sibling's exposure level on the younger sibling's ADHD symptoms.

For the first case, the younger sibling's exposure level was regressed on the older siblings ADHD symptoms while adjusting for the older sibling's exposure level. Results are shown in Table S11.

For the second case, families were stratified based on which sibling (older or younger) had the higher exposure level. Then, sibling analyses were conducted in the two groups of families and effect estimates compared. Results are shown in Tables S12 and S13.

*Assessing the impact of non-shared confounders:*

In sibling control analyses, the within-family effect estimate can be biased if potential confounders are less correlated between siblings (i.e., non-shared) than is the exposure of interest. To determine whether non-shared confounders might impact the observed results, the between-sibling correlations of the exposures of interest were compared to the between-sibling correlations of potential confounders. These results are presented in Tables S14 and S15.

RESULTS:

*Sensitivity analyses: Sex differences*

Sex differences were examined by repeating the sibling control analyses in the ABCD cohort among families with male-male sibling pairs (N=200 families) and female-female sibling pairs (N=176 families) separately. Results for the male-only analyses align very closely to the primary analyses. Significant between-family effects were observed for maternal health, infant health, and birth characteristics, but no within-family effects were significant (Supplemental Table S7). In contrast, in the female-only analyses there were no significant between-family effects, but there was a significant within-family effect for prenatal substance exposures ($\beta$=0.299, p=2.76e-03) (Supplemental Table S8). Due to low sample sizes, we were unable perform sex-stratified analyses in the Oregon ADHD-1000.

*Sensitivity analyses: Examining sources of bias in sibling-control studies*

In addition to conducting models that adjusted for measured confounders, several potential sources of bias in sibling analyses were investigated. Because we did not find statistically significant within-family effects in the sibling analyses, we first focus on the impact of measurement error, which can attenuate these effects. Simulations conducted with the SibSim tool (Gustavson et al., 2024) were used to examine the risk of false-negative findings (i.e., falsely concluding that confounding exists when there is a true causal effect) in the sibling analyses due to measurement error for the exposures studied. We assessed the effect of measurement error in three ways: (1) the amount of attenuation of a true causal effect due to modest measurement error (reliability=0.8), (2) the risk of a false-negative finding, and (3) the power to detect a true causal effect (within-family effect). The details of these simulations are presented in the Supplemental Materials.

Results indicated only a small potential impact of measurement error in our analyses. For nearly all exposures examined in the ABCD cohort, the reduction in effect size between the uncontrolled model (full cohort; Table 3) and the sibling-control model (Table 4) was larger than simulations suggest would be expected due to modest measurement error (reliability=0.8) alone. The one exception is for prenatal substance exposures. In that case, the simulated reduction in effect size, due to measurement error alone, between an uncontrolled model and a sibling-control model was 75%. In the ABCD cohort (Tables 3 and 4), we observed an effect size reduction of 66%, indicating this could be due to measurement error alone and not due to confounding. It should also be noted that family conflict and the stressful life events score both had observed between-sibling correlations above 0.8, and therefore simulations could not be run with measurement reliability of 0.8. Instead, for these two exposures, we report results using a reliability of 0.9. The strong between-sibling correlations for these exposures also substantially reduces the power to detect within-family effects. Results for all simulations are shown in Supplemental Tables S9 and S10.

Given a measurement reliability of 0.8 and using p=0.007 as the threshold for a significant between-family effect, the risk of falsely claiming the existence of confounding with family-level factors ranged from 32.4% for prenatal substance exposures to <1% for infant health problems. In the worst-case scenario, when exposure reliability equals the observed sibling correlation, the risk of false-negative effects ranged from 52.2% for prenatal substance exposure to 13.4% for breastfeeding duration. It should be noted that p-values for between-family effects for both maternal health and infant health factors in ABCD were orders of magnitude smaller (Supplemental Table S4) than the 0.007 threshold used in these simulations.

Second, there was some evidence of carry-over effects impacting the observed results. In the ABCD cohort, there were significant effects of the older sibling's ADHD symptoms on the maternal health factor of the younger sibling ($\beta$=0.127, p=3.15e-4) and the birth characteristics of the younger sibling ($\beta$=0.107, p=0.005). Carry-over effects on family conflict and infant health problems were nominally significant (Supplemental Table S11). However, it is unlikely that the magnitude of these carry-over effects would lead to the amount of effect attenuation observed between the full-cohort analyses and the sibling-control analyses (Tables 3 and 4) (Sjölander et al., 2016).

Results of stratified analyses examining carry-over from the older sibling's exposure to the younger sibling's ADHD symptoms do not indicate the presence of carry-over effects—no statistically significant within family effects are seen in either group of families (Supplemental Tables S12 and S13). However, in the ABCD cohort, there is a nominally significant inverse effect for stressful life events among families where the older child had the higher exposure, suggesting potential long-term impacts from stress in the family environment.

Finally, the risk of non-shared confounders impacting the observed results, in either cohort, appears low. Between sibling correlations of selected observed confounders were stronger than nearly all between sibling correlations of the exposures tested (Supplemental Tables S14 and S15). The one exception was the family conflict measure in the ABCD cohort, which had a between-sibling correlation of 0.89. This is slightly higher than the between-sibling correlation of parental GAF (0.87).

TABLES:

| | ABCD Matched ADHD Case-Control Sub-sample |
|---|---|
| Participants | 1231 (411 ADHD, 820 non-ADHD) |
| Age in years | 9.43 (0.506) |
| % Female | 46.2% |
| Race / Ethnicity | |
| White, non-Hispanic | 621 |
| Hispanic / Latino | 228 |
| Black / African-American | 207 |
| Asian | 6 |
| Other / Unknown | 169 |
| ADHD T-score* | 57.1 (9.26) |
| Family Income | Median = $75,000 - $99,999 |
| Family Conflict | 2.74 (2.05) |
| Parental Stress | 3.43 (2.65) PLE scale |
| Maternal Health Score | 0.116 (0.700) |
| Prenatal Exposure Score | 0.192 (0.740) |
| Infant Health Score | 0.068 (0.640) |
| Birth Characteristics Score | -0.004 (0.790) |
| Breastfeeding duration | 7.38 (8.17) months |

Table S1. Overview of matched ADHD case-control sub-sample of ABCD. *ADHD symptoms were measured by the CBCL ADHD DSM-oriented T-score.

| | ABCD Matched Sub-sample | | ABCD Matched Sub-sample (Adjusted) | |
|---|---|---|---|---|
| | β (SE) | P-value (N) | β (SE) | P-value (N) |
| **Family Conflict** | **0.465 (0.043)** | **1.48e-25 (1231)** | **0.289 (0.046)** | **6.22e-10 (1055)** |
| **Stressful Events** | **0.401 (0.047)** | **5.29e-17 (1159)** | **0.205 (0.050)** | **4.46e-05 (994)** |
| **Maternal Health Prob.** | **0.327 (0.046)** | **1.36e-12 (1224)** | **0.149 (0.047)** | **0.002 (1055)** |
| Prenatal Exposures | **0.289 (0.042)** | **1.50e-11 (1224)** | 0.107 (0.046) | 0.018 (1055) |
| Infant Health Prob. | **0.215 (0.049)** | **1.34e-05 (1224)** | 0.125 (0.048) | 0.010 (1055) |
| Birth Characteristics | **0.164 (0.050)** | **9.66e-04 (1224)** | 0.081 (0.048) | 0.092 (1055) |
| **Breastfeeding Dur.** | **-0.180 (0.053)** | **7.28e-04 (1187)** | **-0.151 (0.051)** | **0.003 (1036)** |

Table S2. Associations in matched ADHD case-control sub-sample of ABCD. Exposures in bold were significant after accounting for multiple testing. Regression models included covariates for age, sex, and the first three genomic principal components, as well as random effects for family and study site. Adjusted models (right-hand column) included additional covariates for child polygenic risk for ADHD, maternal age at birth, highest parental education, household income, parent ADHD symptoms, parent depression problems, and parent GAF score.

| | ADHDRS Total Symptoms T-score | | Conners Inattention Raw Score | |
|---|---|---|---|---|
| | β (SE) | P-value (N) | β (SE) | P-value (N) |
| **Family Conflict** | **0.171 (0.037)** | **5.65e-6 (719)** | **0.132 (0.037)** | **0.000386 (720)** |
| Stressful Events | 0.101 (0.040) | 0.013 (646) | 0.077 (0.040) | 0.054 (646) |
| **Maternal Health Prob.** | **0.215 (0.037)** | **6.17e-9 (738)** | **0.180 (0.036)** | **7.56e-7 (739)** |
| **Prenatal Exposures** | **0.219 (0.037)** | **3.64e-9 (738)** | **0.176 (0.036)** | **1.39e-6 (739)** |
| **Infant Health Prob.** | **0.184 (0.037)** | **6.08e-7 (738)** | **0.143 (0.036)** | **8.76e-5 (739)** |
| Birth Characteristics | 0.057 (0.037) | 0.129 (738) | 0.038 (0.037) | 0.299 (739) |
| **Breastfeeding Dur.** | **-0.188 (0.037)** | **5.80e-7 (734)** | **-0.189 (0.037)** | **2.89e-7 (735)** |

Table S3. Associations with additional ADHD measures available in the Oregon ADHD-1000. Regression models included covariates for age, sex, and the first three genomic principal components, as well as a random effect for family.

| | Within-family Effect | | Between-family Effect | |
|---|---|---|---|---|
| | β (SE) | P-value (N) | β (SE) | P-value (N) |
| Family Conflict | 0.033 (0.074) | 0.654 (553) | 0.234 (0.090) | 0.010 (553) |
| Stressful Events | -0.044 (0.078) | 0.572 (962) | 0.190 (0.086) | 0.027 (962) |
| Maternal Health Prob. | -0.046 (0.044) | 0.293 (1525) | **0.264 (0.054)** | **1.23e-06 (1525)** |
| Prenatal Exposures | 0.059 (0.061) | 0.335 (1523) | 0.097 (0.067) | 0.150 (1523) |
| Infant Health Prob. | -0.014 (0.038) | 0.716 (1523) | **0.203 (0.051)** | **8.05e-05 (1523)** |
| Birth Characteristics | -0.018 (0.042) | 0.674 (1523) | 0.133 (0.054) | 0.014 (1523) |
| Breastfeeding Dur. | 0.020 (0.058) | 0.725 (961) | -0.103 (0.071) | 0.146 (961) |

Table S4. Within- and between-family effects in ABCD. Regression models included covariates for age and sex, as well as a random effect for family.

| | Within-family Effect | | Between-family Effect | |
|---|---|---|---|---|
| | β (SE) | P-value (N) | β (SE) | P-value (N) |
| Family Conflict | 0.162 (0.128) | 0.207 (190) | 0.042 (0.154) | 0.786 (190) |
| Stress | -- | -- | -- | -- |
| Maternal Health Prob. | 0.191 (0.119) | 0.111 (286) | -0.026 (0.137) | 0.850 (286) |
| Prenatal Exposures | 0.236 (0.118) | 0.046 (284) | -0.092 (0.136) | 0.502 (284) |
| Infant Health Prob. | 0.154 (0.104) | 0.140 (288) | -0.054 (0.126) | 0.669 (288) |
| Birth Characteristics | -- | -- | -- | -- |
| Breastfeeding Dur. | -0.082 (0.160) | 0.610 (123) | -0.222 (0.195) | 0.256 (123) |

Table S5. Within- and between-family effects in Oregon ADHD-1000. Regression models included covariates for age and sex, as well as a random effect for family.

| | ABCD | | Oregon ADHD-1000 | |
|---|---|---|---|---|
| | β (SE) | P-value (N) | β (SE) | P-value (N) |
| **Family Conflict** | **0.173 (0.010)** | **2.69e-64 (9729)** | **0.142 (0.040)** | **3.67e-04 (641)** |
| Stress | 0.123 (0.010) | 2.63e-33 (9729) | 0.066 (0.040) | 0.102 (641) |
| **Maternal Health Prob.** | **0.107 (0.010)** | **2.52e-25 (9691)** | **0.165 (0.039)** | **2.95e-05 (637)** |
| **Prenatal Exposures** | **0.127 (0.010)** | **2.34e-35 (9691)** | **0.157 (0.039)** | **7.49e-05 (637)** |
| **Infant Health Prob.** | **0.059 (0.010)** | **5.51e-09 (9691)** | **0.133 (0.039)** | **7.98e-04 (637)** |
| Birth Characteristics | 0.038 (0.011) | 3.23e-04 (9691) | 0.040 (0.039) | 0.307 (637) |
| **Breastfeeding Dur.** | **-0.049 (0.010)** | **2.57e-06 (9391)** | **-0.186 (0.039)** | **2.63e-06 (633)** |

Table S6. Full-cohort analyses adjusted for potential confounders related to trauma and community environment. Exposures in bold were statistically significant in both cohorts. Regression models included covariates for age, sex, the first three genomic principal components, family conflict/stressful events (when not the variable of interest), child opportunity index scales (education, health/environment, social/economic, and total score), and the area deprivation index. Models also included random effects for family and study site (ABCD only).

| | Within-family Effect | | Between-family Effect | |
|---|---|---|---|---|
| | β (SE) | P-value (N) | β (SE) | P-value (N) |
| Family Conflict | 0.020 (0.137) | 0.885 (163) | 0.282 (0.167) | 0.0945 (163) |
| Stressful Events | -0.094 (0.134) | 0.482 (249) | 0.305 (0.154) | 0.0493 (249) |
| Maternal Health Prob. | -0.132 (0.084) | 0.117 (409) | **0.427 (0.104)** | **5.27e-5 (409)** |
| Prenatal Exposures | -0.053 (0.118) | 0.657 (409) | 0.179 (0.132) | 0.177 (409) |
| Infant Health Prob. | -0.066 (0.069) | 0.343 (409) | **0.400 (0.097)** | **4.29e-5 (409)** |
| Birth Characteristics | -0.079 (0.082) | 0.341 (409) | **0.304 (0.104)** | **3.87e-3 (409)** |
| Breastfeeding Dur. | -0.056 (0.102) | 0.586 (254) | -0.079 (0.132) | 0.550 (254) |

Table S7. Within- and between-family effects in ABCD among families with male-male sibling pairs only. Effects in bold were significant after accounting for multiple testing. Regression models were the same as those reported in Table S4.

| | Within-family Effect | | Between-family Effect | |
|---|---|---|---|---|
| | β (SE) | P-value (N) | β (SE) | P-value (N) |
| Family Conflict | 0.150 (0.132) | 0.257 (139) | 0.121 (0.172) | 0.480 (139) |
| Stressful Events | -0.014 (0.137) | 0.920 (232) | 0.0891 (0.160) | 0.578 (232) |
| Maternal Health Prob. | 0.115 (0.086) | 0.182 (367) | 0.0493 (0.110) | 0.654 (367) |
| Prenatal Exposures | **0.299 (0.099)** | **0.00276 (365)** | -0.092 (0.117) | 0.434 (365) |
| Infant Health Prob. | 0.075 (0.770) | 0.332 (367) | -0.019 (0.106) | 0.857 (367) |
| Birth Characteristics | 0.039 (0.081) | 0.632 (365) | -0.038 (0.109) | 0.730 (365) |
| Breastfeeding Dur. | 0.122 (0.121) | 0.317 (229) | -0.117 (0.148) | 0.432 (229) |

Table S8. Within- and between-family effects in ABCD among families with female-female sibling pairs only. Exposures in bold were significant after accounting for multiple testing. Regression models are the same as those reported in Table S4.

| **Exposure** | **Causal Effect Size** | **Observed Correlation** | **Reliability** | **Uncontrolled Effect Est.** | **Within-family Est.** | **% Reduction** |
|---|---|---|---|---|---|---|
| Family Conflict | 0.2 | 0.89 | 0.95 | 0.18 | 0.11 | 39% |
| | 0.2 | 0.89 | 0.9 | 0.16 | 0.03 | 81% |
| | 0.2 | 0.89 | 0.89 | 0.16 | 0.01 | 94% |
| Stress | 0.16 | 0.84 | 0.95 | 0.15 | 0.11 | 27% |
| | 0.16 | 0.84 | 0.9 | 0.13 | 0.06 | 54% |
| | 0.16 | 0.84 | 0.84 | 0.12 | 0.01 | 92% |
| Maternal Health | 0.15 | 0.51 | 0.9 | 0.13 | 0.12 | 8% |
| | 0.15 | 0.51 | 0.8 | 0.11 | 0.09 | 18% |
| | 0.15 | 0.51 | 0.7 | 0.09 | 0.06 | 33% |
| Prenatal Exposures | 0.18 | 0.76 | 0.9 | 0.15 | 0.11 | 27% |
| | **0.18** | **0.76** | **0.8** | **0.12** | **0.03** | **75%** |
| | 0.18 | 0.76 | 0.76 | 0.11 | 0.01 | 91% |
| Infant Health | 0.1 | 0.38 | 0.9 | 0.09 | 0.08 | 11% |
| | 0.1 | 0.38 | 0.8 | 0.08 | 0.07 | 13% |
| | 0.1 | 0.38 | 0.7 | 0.06 | 0.05 | 17% |
| Birth | 0.1 | 0.50 | 0.9 | 0.09 | 0.08 | 11% |
| | 0.1 | 0.50 | 0.8 | 0.07 | 0.06 | 14% |
| | 0.1 | 0.50 | 0.7 | 0.06 | 0.04 | 33% |

| | | | | | | |
|---|---|---|---|---|---|---|
| Breast-feeding Duration | 0.1 | 0.68 | 0.9 | 0.09 | 0.07 | 22% |
| | 0.1 | 0.68 | 0.8 | 0.07 | 0.04 | 43% |
| | 0.1 | 0.68 | 0.7 | 0.06 | 0.01 | 83% |

Table S9. Effect estimate attenuation due to measurement error. For all scenarios, the number of families was set to 700, the percentage of families with more than two siblings was set at 1%, and 500 random samples were performed. In all cases, the outcome was treated as continuous.

| Exposure | Causal Effect Size | Observed Correlation | Reliability | Risk of False-negative | Power |
|---|---|---|---|---|---|
| Family Conflict | 0.2 | 0.89 | 0.95 | 6.4% | 11.6% |
| | 0.2 | 0.89 | 0.9 | 25.4% | 1.6% |
| | 0.2 | 0.89 | 0.89 | 34% | 0.8% |
| Stress | 0.16 | 0.84 | 0.95 | 1.4% | 18.4% |
| | 0.16 | 0.84 | 0.9 | 10% | 5.8% |
| | 0.16 | 0.84 | 0.84 | 27.2% | 0.8% |
| Maternal Health | 0.15 | 0.51 | 0.9 | 1% | 75.6% |
| | 0.15 | 0.51 | 0.8 | 3% | 49.8% |
| | 0.15 | 0.51 | 0.7 | 10% | 21.8% |
| | 0.15 | 0.51 | 0.51 | 47.4% | 0.8% |
| Prenatal Exposures | 0.18 | 0.76 | 0.9 | 6.6% | 30% |
| | 0.18 | 0.76 | 0.8 | 32.4% | 2.4% |
| | 0.18 | 0.76 | 0.76 | 52.2% | 0.8% |
| Infant Health | 0.1 | 0.38 | 0.9 | 0.4% | 47.8% |
| | 0.1 | 0.38 | 0.8 | 0.6% | 36.4% |
| | 0.1 | 0.38 | 0.7 | 1.8% | 19.6% |
| | 0.1 | 0.38 | 0.38 | 16% | 0.8% |
| Birth | 0.1 | 0.50 | 0.9 | 0.6% | 33.8% |

| | | | | | |
|---|---|---|---|---|---|
| | 0.1 | 0.50 | 0.8 | 1.4% | 19.2% |
| | 0.1 | 0.50 | 0.7 | 3.4% | 9.2% |
| | 0.1 | 0.50 | 0.5 | 16.4% | 0.8% |
| Breast-feeding Duration | 0.1 | 0.68 | 0.9 | 1% | 13.8% |
| | 0.1 | 0.68 | 0.8 | 4.8% | 5.6% |
| | 0.1 | 0.68 | 0.7 | 12% | 1.4% |
| | 0.1 | 0.68 | 0.68 | 13.4% | 0.8% |

Table S10. Risk of a false-negative finding and power to detect a true causal (within-family) effect.

| | ABCD | | Oregon ADHD-1000 | |
|---|---|---|---|---|
| **Exposure** | **β (SE)** | **P-value (N)** | **β (SE)** | **P-value** |
| Family Conflict | 0.121 (0.059) | 0.041 (245) | 0.008 (0.107) | 0.937 (90) |
| Stress | 0.033 (0.036) | 0.357 (436) | 0.222 (0.140) | 0.120 (52) |
| **Maternal Health Prob.** | **0.127 (0.035)** | **3.15e-04 (691)** | 0.017 (0.069) | 0.807 (133) |
| Prenatal Exposures | 0.031 (0.026) | 0.240 (690) | -0.013 (0.071) | 0.857 (132) |
| Infant Health Prob. | 0.091 (0.038) | 0.018 (690) | -0.023 (0.072) | 0.746 (134) |
| **Birth Characteristics** | **0.107 (0.038)** | **0.005 (690)** | -0.010 (0.074) | 0.892 (134) |
| Breastfeeding Dur. | 0.020 (0.044) | 0.644 (434) | 0.109 (0.137) | 0.427 (59) |

Table S11. Assessing carry-over effects of the older sibling's ADHD symptoms on the younger sibling's exposure level. Here, N = number of families, not number of subjects. Effects in bold were significant after accounting for multiple testing. In regression models, the younger sibling's exposure level was the dependent variable and the older sibling's ADHD symptom score was the independent variable. The older sibling's exposure level was covaried.

| | Families in which the <u>older</u> sibling has higher exposure | | Families in which the <u>younger</u> sibling has the higher exposure | |
|---|---|---|---|---|
| **Exposure** | **β (SE)** | **P-value (N)** | **β (SE)** | **P-value (N)** |
| Family Conflict | 0.198 (0.145) | 0.174 (292) | -0.194 (0.144) | 0.180 (198) |
| Stress | -0.287 (0.135) | 0.035 (540) | 0.045 (0.163) | 0.782 (332) |
| Maternal Health Prob. | -0.021 (0.073) | 0.778 (782) | -0.043 (0.084) | 0.614 (600) |
| Prenatal Exposures | 0.073 (0.090) | 0.420 (768) | 0.215 (0.131) | 0.102 (612) |
| Infant Health Prob. | -0.113 (0.068) | 0.097 (762) | 0.060 (0.066) | 0.365 (618) |
| Birth Characteristics | -0.025 (0.076) | 0.744 (730) | -0.032 (0.072) | 0.655 (650) |
| Breastfeeding Dur. | 0.099 (0.106) | 0.352 (406) | 0.057 (0.100) | 0.568 (462) |

Table S12. Assessing carry-over effects of the older-sibling's exposure level on the younger sibling's ADHD symptoms in ABCD. Estimates of the within-family effect of each exposure on ADHD symptoms are shown. Regression models were the same as those reported in Table S4.

| | Families in which the <u>older</u> sibling has higher exposure | | Families in which the <u>younger</u> sibling has the higher exposure | |
|---|---|---|---|---|
| **Exposure** | **β (SE)** | **P-value (N)** | **β (SE)** | **P-value (N)** |
| Family Conflict | 0.298 (0.203) | 0.147 (102) | 0.068 (0.263) | 0.797 (78) |
| Stress | 0.397 (0.230) | 0.091 (54) | 0.324 (0.263) | 0.224 (48) |
| Maternal Health Prob. | 0.098 (0.165) | 0.552 (160) | 0.125 (0.260) | 0.632 (106) |
| Prenatal Exposures | 0.201 (0.157) | 0.204 (158) | 0.221 (0.255) | 0.389 (106) |
| Infant Health Prob. | 0.105 (0.140) | 0.453 (160) | 0.057 (0.241) | 0.814 (108) |
| Birth Characteristics | 0.175 (0.166) | 0.295 (144) | -0.071 (0.193) | 0.715 (124) |
| Breastfeeding Dur. | 0.053 (0.266) | 0.842 (64) | -0.162 (0.333) | 0.630 (54) |

Table S13. Assessing carry-over effects of the older-sibling's exposure level on the younger sibling's ADHD symptoms in the Oregon ADHD-1000. Within-family effect estimates are shown. Regression models were the same as those reported in Table S5.

| **Exposure** | **ABCD** | **Oregon ADHD-1000** |
|---|---|---|
| Family Conflict | 0.895 | 0.520 |
| Stress | 0.837 | 0.448 |
| Maternal Health Prob. | 0.505 | 0.653 |
| Prenatal Exposures | 0.755 | 0.631 |
| Infant Health Prob. | 0.384 | 0.558 |
| Birth Characteristics | 0.500 | 0.717 |
| Breastfeeding Dur. | 0.675 | 0.689 |

Table S14. Between-sibling correlations for the exposures examined.

| **Potential Confounder** | **ABCD** | **Oregon ADHD-1000** |
|---|---|---|
| Parental Education | 0.982 | 0.755 |
| Family Income | 0.984 | 0.912 |
| Parental ADHD | 0.921 | 1.0 |
| Parental MDD | 0.971 | 0.975 |
| Parental GAF | 0.873 | 0.906 |
| COI | 0.979 | 0.912 |
| ADI | 0.986 | 0.896 |

Table S15. Observed between-sibling correlations for potential family-level confounders. COI = child opportunity index; ADI = area deprivation index. Tetrachoric correlations were calculated for dichotomous variables (parental MDD in ABCD; parental ADHD and MDD in Oregon ADHD-1000).
